# Estimating historical disease burden and the impact of vaccination by influenza type and subtype in the United States, 2016-2020

**DOI:** 10.1101/2024.10.25.24316040

**Authors:** Sinead E. Morris, Sarabeth M. Mathis, Jessie R. Chung, Brendan Flannery, Alissa O’Halloran, Charisse N. Cummings, Shikha Garg, Peng-Jun Lu, Tammy A. Santibanez, Carrie Reed, Matthew Biggerstaff, A. Danielle Iuliano

## Abstract

Seasonal influenza causes substantial morbidity and mortality in the United States. The U.S. Centers for Disease Control and Prevention (CDC) uses a compartmental framework to estimate the annual disease burden and burden prevented by vaccination for all influenza types and subtypes combined. However, these estimates do not capture underlying shifts in disease burden caused by different circulating influenza virus types or subtypes. We demonstrate an extension of the current framework to estimate disease burden and burden prevented by vaccination for influenza A virus subtypes A(H1N1) and A(H3N2), and influenza type B viruses. We applied this method to data from the 2016/17 to 2019/20 seasons that include age- and virus-specific hospitalizations and vaccine effectiveness estimates, and age-specific vaccination coverage estimates. We estimated the number of symptomatic illnesses, medically-attended illnesses, hospitalizations, and deaths caused by each virus, and the corresponding number prevented by vaccination. Disease burden and vaccine-prevented burden varied substantially by season, age, and virus type or subtype. The greatest disease burden was estimated in 2017/18, whereas 2019/20 had the greatest burden prevented by vaccination. Influenza A viruses contributed most to disease burden in all seasons. Vaccination against influenza B viruses prevented the largest percentage of hospitalizations among children and adults <65 years, whereas vaccination against A(H1N1) prevented the largest percentage of hospitalizations among adults ≥65 years. Overall, our results highlight complex variability in influenza disease burden by season, age, and virus type and subtype. These findings can be used to improve understanding of the factors impacting influenza disease burden each season and to enhance communications of the value of influenza vaccination.

**Highlights:**

- Estimates of disease burden and vaccine-prevented burden inform influenza guidance.
- Contributions to burden from each virus type and subtype vary by season and age.
- A(H3N2) caused the greatest total disease burden from the 2016/17 to 2019/20 seasons.
- Children <18 years experienced greater influenza B burden compared with other ages.
- Vaccination against A(H1N1) and B prevented the greatest percentage of severe disease.

## Introduction

Seasonal influenza viruses cause substantial morbidity and mortality in the United States. The U.S. Centers for Disease Control and Prevention (CDC) estimates that between 9–41 million symptomatic illnesses, 100,000–710,000 hospitalizations, and 5,000–52,000 deaths occur each year due to influenza^1^. Seasonal influenza vaccination is recommended for all persons ≥6 months old without contraindications and prevents an estimated 1–7.5 million symptomatic illnesses, 22,000–100,000 hospitalizations and 1,000–12,000 deaths annually^2^. Estimating influenza disease burden and the burden prevented by vaccination is important for assessing the severity of seasonal influenza epidemics and communicating the benefits of vaccination.

Currently, CDC estimates burden and burden prevented by vaccination using a compartmental framework that models all influenza virus types and subtypes combined^3,4^. However, these estimates do not capture underlying shifts in disease burden caused by different influenza virus types (A vs. B) and different influenza A subtypes (A(H1N1) vs. A(H3N2)) that can change seasonally due to complex interactions between virus evolution, population immunity, and vaccine effectiveness (VE). Estimating the different contributions of each type or subtype to overall disease burden and burden prevented by vaccination could improve our understanding of the impact of influenza each season and its changing epidemiological landscape.

In this paper, we demonstrate an extension of the current CDC framework to estimate disease burden and burden prevented by vaccination by influenza virus type and subtype. We applied this method to data from the 2016/17 to 2019/20 seasons that include age- and virus-specific hospitalizations and VE estimates, and age-specific vaccination coverage estimates. Our method can be combined with annual disease burden calculations to enhance communications of the risks of influenza and the benefits of seasonal influenza vaccination.

## Methods

### Surveillance data

We used surveillance data spanning the 2016/17 to 2019/20 influenza seasons. Unless otherwise specified, data were aggregated into six age groups: 0–6 months, 6 months–4 years, 5–17 years, 18–49 years, 50–64 years, and ≥65 years. Monthly numbers of laboratory-confirmed influenza-associated hospitalizations were obtained from the Influenza Hospitalization Surveillance Network (FluSurv-NET), a population-based hospitalization surveillance network that represents approximately 29 million people in the United States^5^. These data included the total number of hospitalizations, the number of hospitalizations attributed to influenza A and influenza B, and the proportions of influenza A hospitalizations that were attributed to A(H1N1) and A(H3N2) (Tables S1–S2, Figure S1). The latter proportions incorporated imputed influenza A subtype information (which was originally missing for approximately 55-60% of influenza A cases each season)^6,7,8^. FluSurv-NET also provided information on the total catchment population for each age group^9^. To account for under-ascertainment of laboratory confirmed influenza hospitalizations, we calculated age-specific under-detection multipliers based on testing practice information from FluSurv-NET sites and diagnostic test sensitivity^9^.

In addition to the distributions of virus type and subtype among influenza-associated hospitalizations, we obtained corresponding distributions for laboratory-confirmed influenza-associated outpatient visits from the US Influenza Vaccine Effectiveness (Flu VE) Network^10^ (Table S2, Figure S2). Since this system recruits participants for vaccine effectiveness studies, there were no distributions for infants aged 0–6 months (who are not eligible for influenza vaccination). We therefore assumed distributions in that age group were equivalent to distributions among children aged 6 months–4 years. We verified that the distributions from the Flu VE Network and FluSurv-NET were similar to independent data obtained from the World Health Organization (WHO) and National Respiratory and Enteric Virus Surveillance System (NREVSS) Collaborating Labs and reported to CDC’s FluView (Figure S3)^11^.

Finally, vaccine effectiveness estimates for A(H1N1), A(H3N2), influenza B, and all influenza viruses combined were obtained from the Flu VE Network, and vaccine coverage estimates were obtained from FluVaxView for all age groups ≥6 months^12^ (Table S1, Figure S4). We approximated the population size for each age group and season using US Census data^13^.

### Framework for estimating disease burden for influenza A(H1N1), A(H3N2), and B

The methods employed by CDC to estimate disease burden and burden prevented by vaccination for all influenza (i.e., for all viruses combined) have been described previously^3,4,14–17^ (Figure 1A). Briefly, the monthly number of influenza-associated hospitalizations for each age group are first translated to rates by dividing by the FluSurv-NET catchment population. These rates are then corrected for under-detection of influenza-related hospitalizations and extrapolated to estimate total influenza hospitalizations in the US population using the age-specific US census estimates. Numbers of symptomatic influenza illnesses and deaths are obtained by multiplying the extrapolated number of hospitalizations by age-specific ratios for the number of symptomatic influenza cases per hospitalization (case-hospitalization ratio) and deaths per hospitalization (death-hospitalization ratio), respectively^3,18,16^. The number of medically-attended illnesses are obtained by multiplying the number of symptomatic illnesses by age-specific estimates of the fraction of people with influenza-associated illness that seek medical care^19^. Values for the under-detection, case-hospitalization, death-hospitalization, and medically-attended multipliers are provided in Table S3.

**Figure 1.**
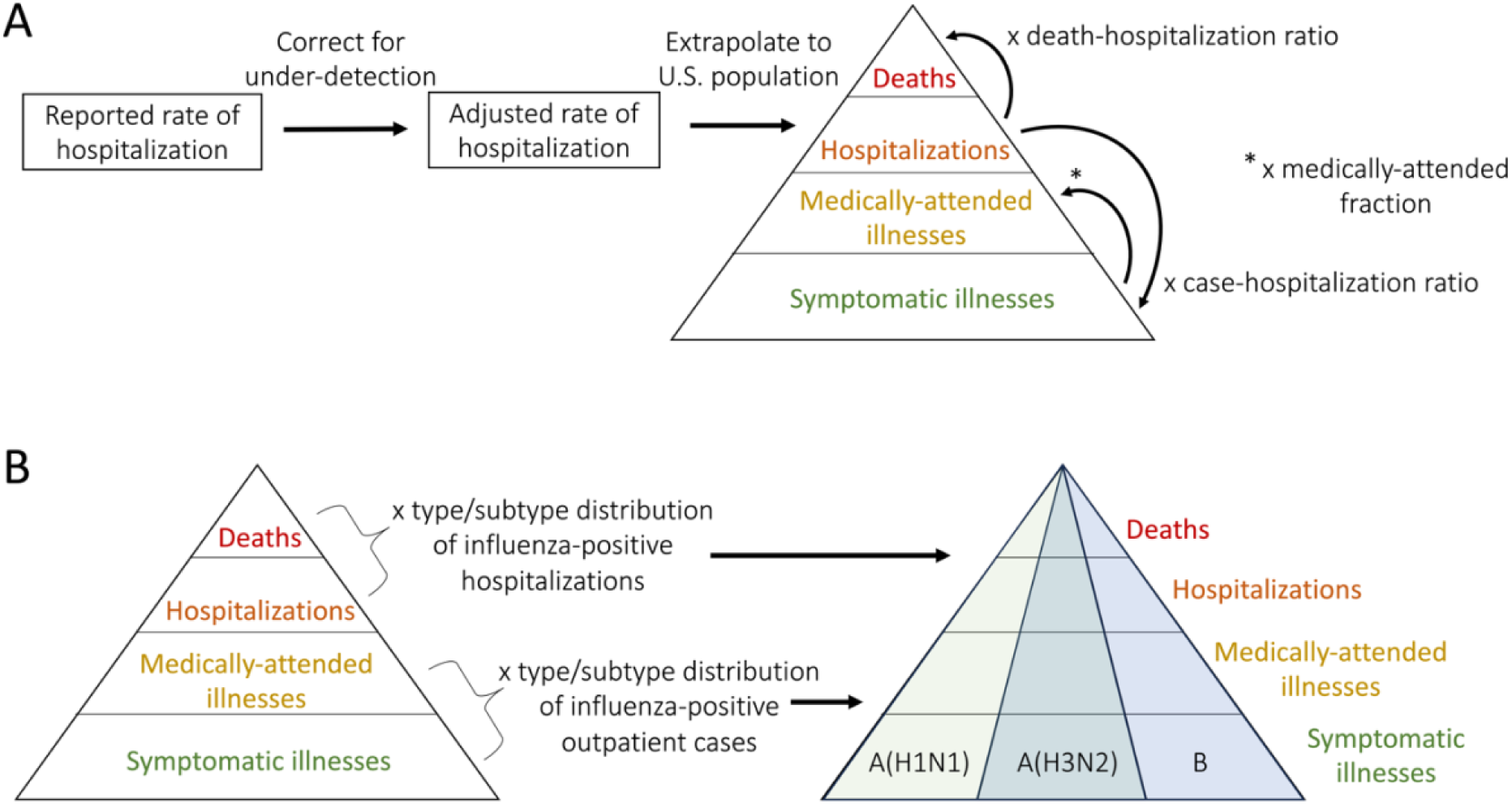
Framework for estimating disease burden. (A) Framework for estimating age-specific burden estimates for all influenza viruses. For each age group, reported hospitalizations rates are corrected for under-detection and extrapolated to the total population. These are then translated to estimates of deaths and symptomatic illnesses using age-specific death-hospitalization ratios and case-hospitalization ratios, respectively. Symptomatic illnesses are translated to medically-attended illnesses using age-specific estimates of the fraction that are medically-attended. (B) Burden estimates are translated to type or subtype specific estimates using virus distributions reported among influenza-positive hospitalizations (from FluSurv-NET) or outpatient cases (from the Flu VE Network).

We extended these methods to generate virus type- and subtype-specific estimates (Figure 1B). For each month and age group, hospitalizations were attributed to either influenza A or B using the relative proportions of each virus type in FluSurv-NET. Hospitalizations attributed to influenza A were further partitioned by subtype using the relative proportions of A(H1N1) and A(H3N2). We used the same type and subtype proportions to partition the total number of deaths into those attributable to A(H1N1), A(H3N2) and B. For symptomatic and medically-attended illnesses, we instead used the relative type and subtype proportions among Flu VE Network outpatient cases as these more accurately reflect virus distributions among non-hospitalized cases of influenza. When presenting results, we combined estimates for the two youngest age groups (0–6 months and 6 months–4 years) into all children aged 0–4 years.

### Framework for estimating burden prevented by vaccination for influenza A(H1N1), A(H3N2) and B

For each virus type or subtype (A(H1N1), A(H3N2), B, or all influenza viruses combined), the monthly, age-specific estimates of disease burden were combined with estimates of vaccine coverage and effectiveness to estimate the burden prevented by vaccination following previously described methods^4^ (Figure 2). We excluded the 0–6 months age group since they are ineligible for vaccination. The population was divided into three categories, susceptible, effectively vaccinated, and (symptomatically) infected as follows. For each age group, *a,* and type or subtype, *i*, the number effectively vaccinated (*EV*) in each successive month *m* is calculated as

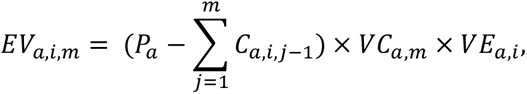

**Figure 2.**
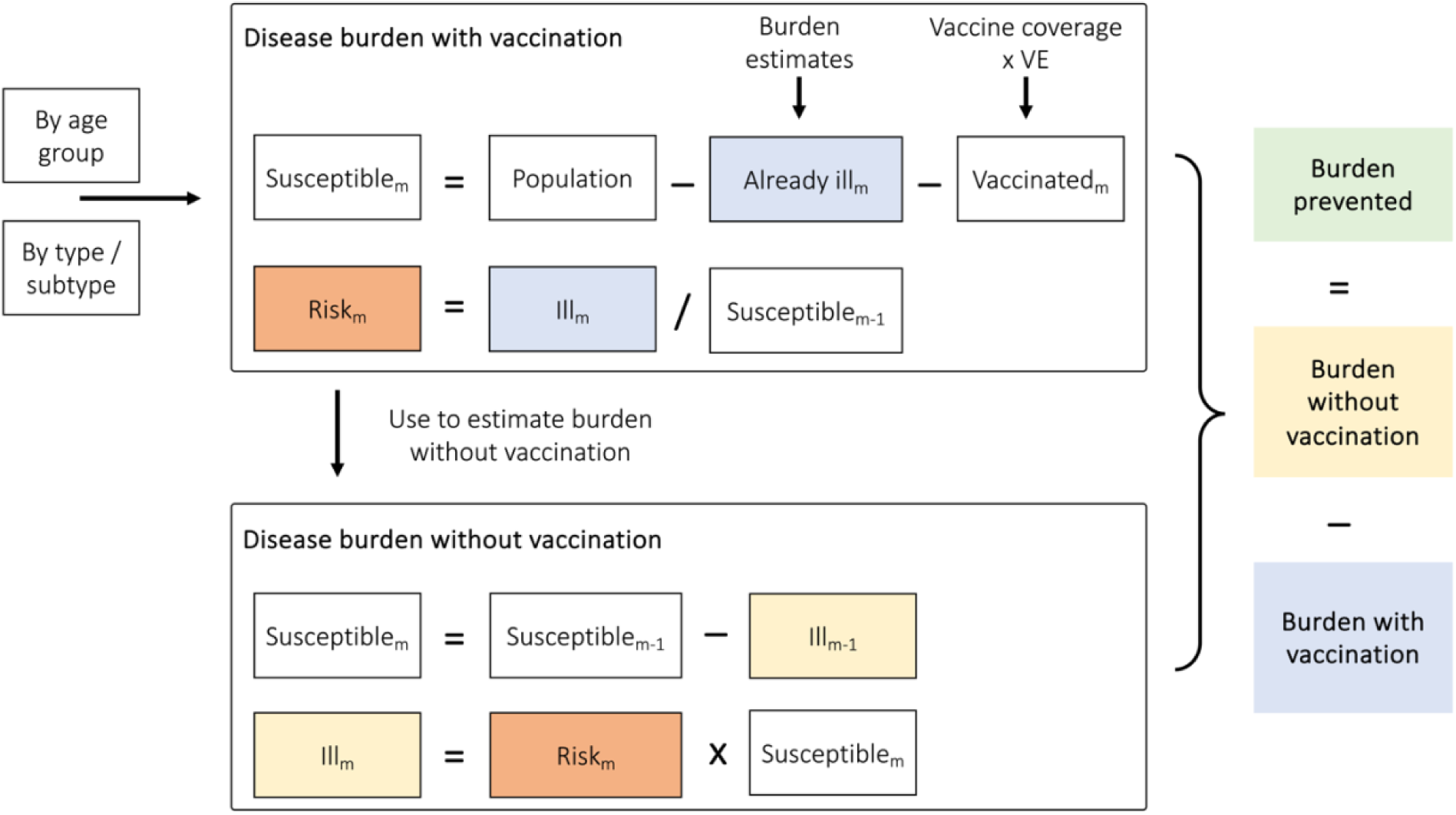
General framework for estimating disease burden prevented by vaccination within each age group and virus type or subtype for a given season. Independent calculations are performed for each age group and virus type or subtype. The procedure is performed iteratively for each month (denoted by the subscript ‘m’). Abbreviations: VE = vaccine effectiveness.

where *P*_*a*_ is the starting population size (assuming everyone is susceptible at the beginning of the season), *VC*_*a*,*m*_ is the proportion vaccinated in that month, *VE*_*a*,*i*_ is the vaccine effectiveness against type or subtype *i*, and 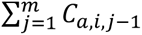 is the cumulative number of symptomatic illnesses estimated during the disease burden calculations and occurring up to the previous month (with *C_a,i, 0_* = 0). This formulation assumes that vaccination provides all-or-nothing protection (i.e., individuals either gain complete immunity following vaccination or remain entirely susceptible) and that susceptible and infected individuals are equally likely to be vaccinated. The number susceptible (S) is then estimated as

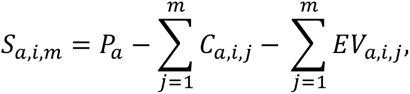

with the implicit assumption that individuals can be infected by more than one virus type or subtype within a season.

The risk of symptomatic illness (*R*) is the number of symptomatic illnesses that occurred in the current month, *m*, divided by the susceptible (i.e., at-risk) population in the previous month, *m–1*,

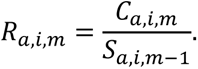

Similarly, the risks of medically-attended illness, hospitalization, and death are approximated by dividing the value in month *m* (estimated during the disease burden calculations) by the susceptible population in month *m–1*. We use these risks to estimate the number of symptomatic illnesses, medically-attended illnesses, hospitalizations, and deaths in the absence of vaccination. The estimated number of symptomatic illnesses without vaccination, *Ĉ*, is

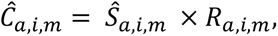

where the susceptible population without vaccination, *Ŝ*, is now calculated iteratively as

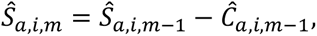

with *Ŝ*_*a*,*i*,0_ = *P*_*a*_ and *Ĉ*_*a*,*i*,0_ = 0. The number of symptomatic illnesses prevented by vaccination is then *Ĉ*_*a*,*i*,*m*_ − *C*_*a*,*i*,*m*_. Analogous calculations were used to estimate the number of medically-attended illnesses, hospitalizations, and deaths prevented by vaccination. For each variable, age group, and type or subtype, we calculated the sum of burden prevented by vaccination across all months in the season. We also calculated disease burden prevented by vaccination as a percentage of the total estimated disease burden in the absence of vaccination, for example, 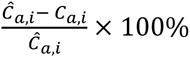. Finally, we note that estimates of burden prevented by vaccination for all influenza viruses combined may not always equal the sum of burden prevented for A(H1N1), A(H3N2), and B viruses because we use independent estimates of VE in each calculation.

### Uncertainty distributions

We generated uncertainty distributions for each disease burden and vaccine-prevented burden estimate using Monte Carlo simulation methods described previously^20,21,22^. Key input variables were each assigned a probability distribution (Table S4), from which we generated 10,000 independent samples. We then re-performed the disease burden and vaccine-prevented burden calculations for each sample and reported uncertainty as the 95^th^ percentiles of the resulting distribution. All analyses were performed in R version 4.2.0^23^ and resulting estimates are reported to three significant figures.

## Results

### Estimated disease burden

For the 2016/17 to 2019/20 seasons, total estimated influenza disease burden ranged from 29.4–41.0 million symptomatic illnesses, 13.6–18.9 million medically-attended illnesses, 395,000–710,000 hospitalizations, and 22,100–51,800 deaths. The greatest number of hospitalizations and deaths occurred in 2017/18 (Figure 3A), and across all seasons the greatest proportion of hospitalizations and deaths occurred among adults ≥65 years (Figure S5).

**Figure 3.**
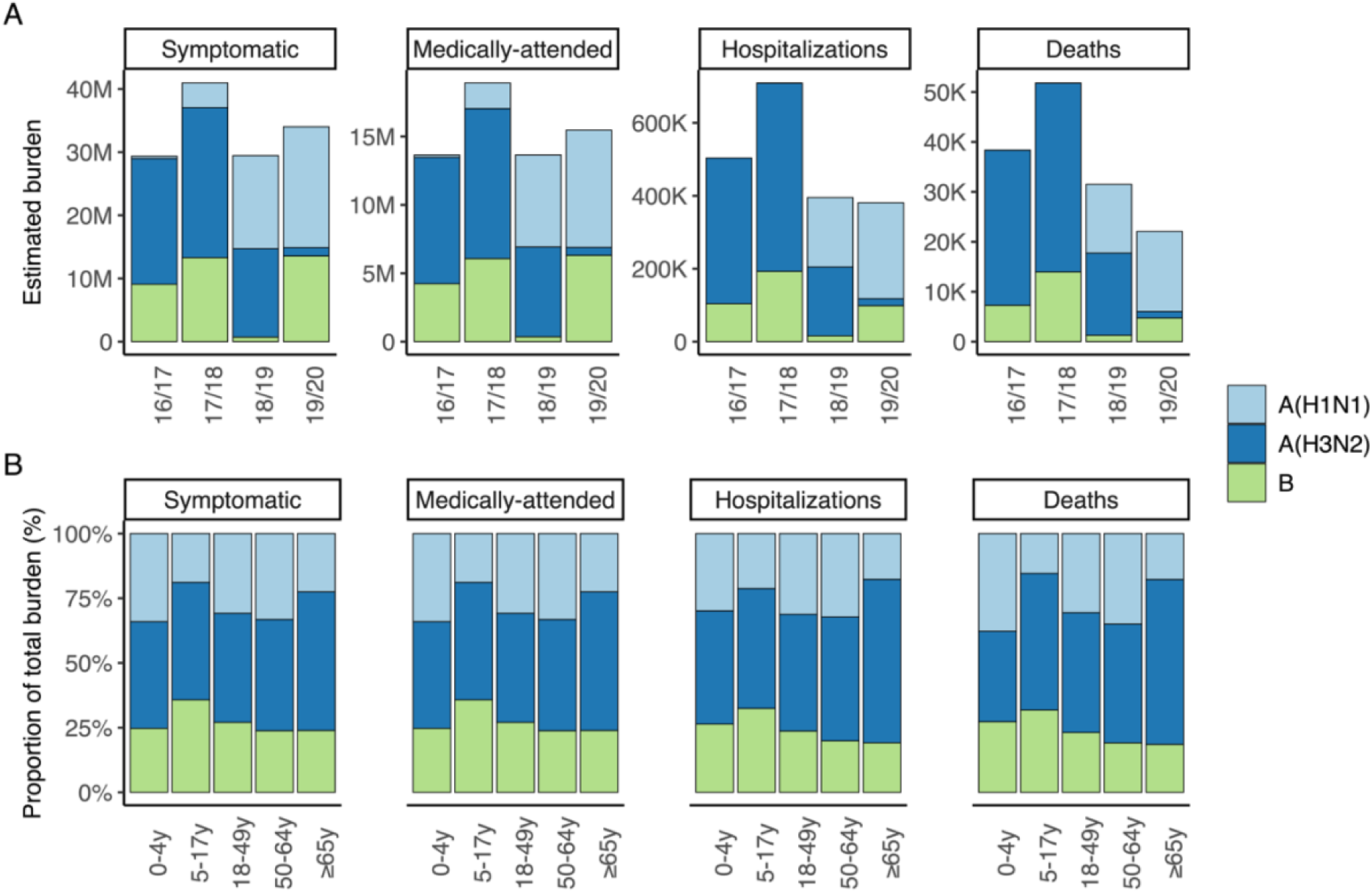
Estimated disease burden. (A) Point estimates of disease burden by type/subtype and season. (B) Distribution of type/subtypes causing burden in each age group, for all four seasons. Estimates by age group, season, and type/subtype, with corresponding 95^th^ percentile uncertainty intervals, are shown in Figure S6.

Among virus types and subtypes, A(H3N2) caused the greatest cumulative burden from 2016/17–2019/20, with 59.0 million total symptomatic illnesses, 27.3 million medically-attended illnesses, 1.12 million hospitalizations and 86,700 deaths (Table 1). However, the contribution of each type and subtype to overall disease burden varied by season and age group (Figure 3, Figure S6). Influenza A(H3N2) and B contributed most burden in 2016/17 and 2017/18, whereas A(H1N1) and A(H3N2) contributed most burden in 2018/19, and A(H1N1) and B contributed most burden in 2019/20 (Figure 3A). For all age groups, influenza A viruses comprised the greatest proportion of burden across all seasons, although the proportion attributed to influenza B viruses increased slightly among children <18 years (Figure 3B).

**Table 1.** Estimated disease burden by virus type or subtype. Numbers represent total estimated burden (and 95^th^ percentile uncertainty intervals) from 2016/17–2019/20 across all age groups and are reported to three significant figures.

| Influenza type or subtype | Symptomatic illnesses (95<sup>th</sup> uncertainty interval) | Medically-attended illnesses (95<sup>th</sup> uncertainty interval) | Hospitalizations (95<sup>th</sup> uncertainty interval) | Deaths (95<sup>th</sup> uncertainty interval) |
| --- | --- | --- | --- | --- |
| A(H1N1) | 38.2 million<br>(34.2–52.8 million) | 17.4 million<br>(15.7–23.4 million) | 456,000<br>(393,000–685,000) | 29,800<br>(20,600–66,500) |
| A(H3N2) | 59.0 million<br>(54.7–72.0 million) | 27.3 million<br>(25.5–33.4 million) | 1.12 million<br>(968,000–1.58 million) | 86,700<br>(63,300–160,000) |
| B | 36.7 million<br>(33.5–48.9 million) | 17.0 million<br>(15.6–22.3 million) | 411,000<br>(365,000–559,000) | 27,200<br>(20,400–48,800) |
| All influenza | 134 million<br>(125–167 million) | 61.7 million<br>(58.0–76.0 million) | 1.99 million<br>(1.76–2.69 million) | 144,000<br>(111,000–256,000) |

### Estimated burden prevented by vaccination

Vaccine coverage was highest among children 6 months–4 years and adults ≥65 years, and during the 2018/19 and 2019/20 seasons (Table S1, Figure S4A). VE varied substantially by season, age group, and virus type or subtype. Broadly, VE was highest in 2017/18 and lowest in 2018/19, decreased with increasing age, and was lower against A(H3N2) than A(H1N1) and B (Table S1, Figure S4B). One notable exception occurred in 2019/20, when adults ≥65 years experienced relatively high VE against A(H1N1) and A(H3N2).

From 2016/17 to 2019/20, total estimated burden prevented by vaccination ranged from 2.85–6.74 million symptomatic illnesses, 1.49–3.28 million medically-attended illnesses, 35,800–98,500 hospitalizations, and 2,520–6,370 deaths. The greatest burden prevented by vaccination occurred in 2019/20 (Figure 4A). For all seasons, the greatest number of hospitalizations were prevented among adults ≥65 years, whereas the greatest percent of hospitalizations were generally prevented among children 6 months–4 years (Figure S7).

**Figure 4.**
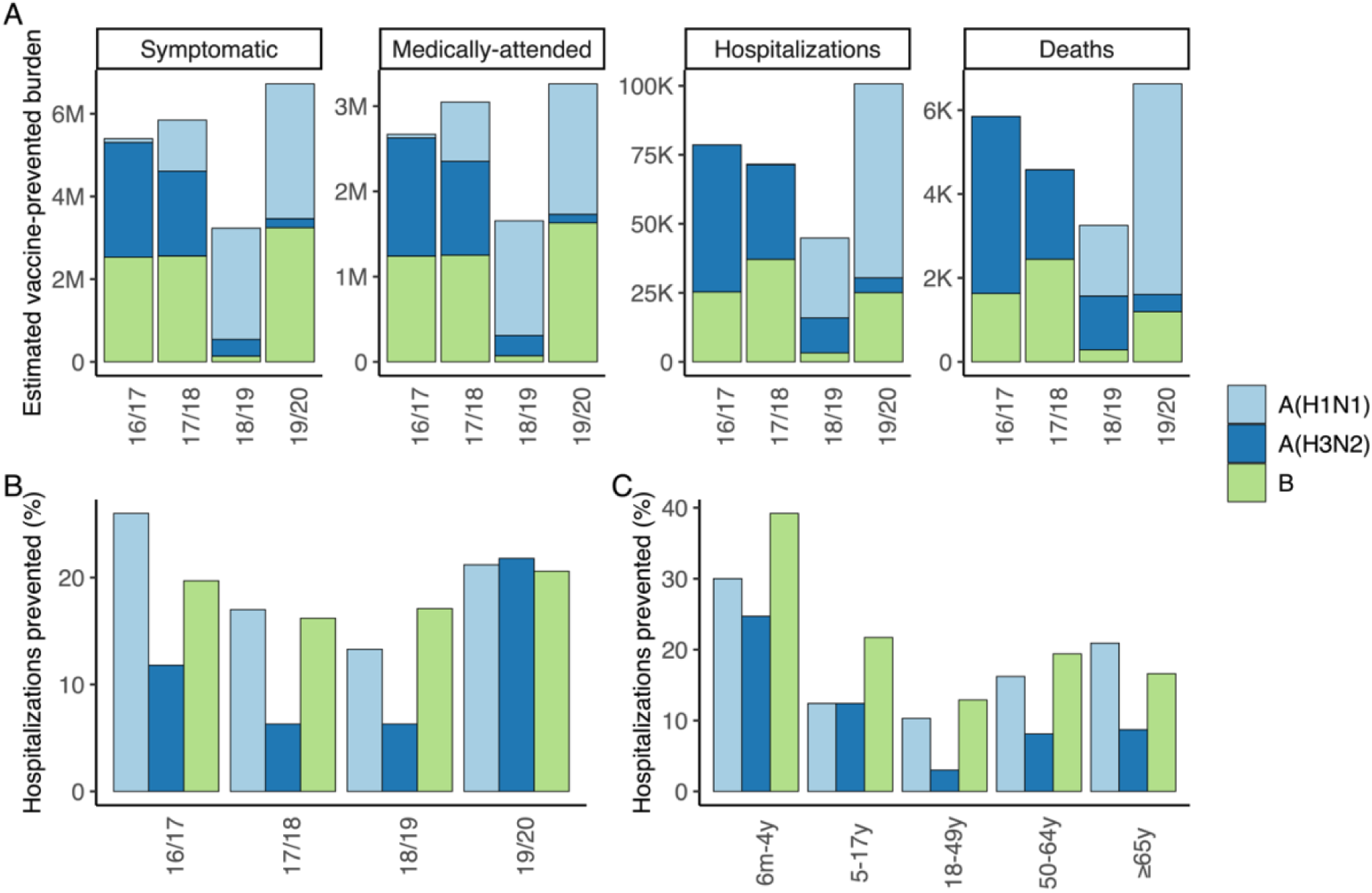
Estimated burden prevented by vaccination. (A) Point estimates for total averted burden by type/subtype and season. (B) Percent of hospitalizations averted by type/subtype and season (for all age groups). (C) Percent of hospitalizations averted by type/subtype and age group (across all four seasons). Estimates by age group, season, and type or subtype, with corresponding 95^th^ percentile uncertainty intervals, are shown in Figure S8.

Among type and subtypes, the largest cumulative number of hospitalizations and deaths were prevented by vaccination against influenza A(H3N2), whereas the largest number of symptomatic and medically-attended illnesses were prevented by vaccination against influenza B (Table 2). In line with our disease burden estimates, vaccination against influenza A(H3N2) and B prevented most burden in 2016/17 and 2017/18, whereas vaccination against A(H1N1) and A(H3N2) prevented most burden in 2018/19, and vaccination against A(H1N1) and B prevented most burden in 2019/20 (Figure 4A, Figure S8). Among seasons, the percent of hospitalizations prevented was generally highest for A(H1N1) and B, with substantially lower percentages for A(H3N2) (except in 2019/20 when the overall burden of A(H3N2) was low) (Figure 4B). This was also true among age groups, with vaccination against influenza B preventing the highest percentage of hospitalizations among children and adults <65 years, and vaccination against A(H1N1) preventing the highest percentage among adults ≥65 years (Figure 4C).

**Table 2.** Estimated vaccine-prevented disease burden by virus type or subtype. Numbers represent total prevented disease burden (and 95^th^ percentile uncertainty intervals) from 2016/17–2019/20 across all age groups and are reported to three significant figures.

| Influenza type or subtype | Symptomatic illnesses (95<sup>th</sup> uncertainty interval) | Medically-attended illnesses (95<sup>th</sup> uncertainty interval) | Hospitalizations (95<sup>th</sup> uncertainty interval) | Deaths (95<sup>th</sup> uncertainty interval) |
| --- | --- | --- | --- | --- |
| A(H1N1) | 7.28 million<br>(6.19–9.83 million) | 3.62 million<br>(3.10–4.81 million) | 99,500<br>(71,100–172,000) | 6,720<br>(3,780–17,100) |
| A(H3N2) | 5.45 million<br>(4.75–7.76 million) | 2.83 million<br>(2.45–4.05 million) | 106,000<br>(75,000–195,000) | 8,050<br>(4,750–19,500) |
| B | 8.47 million<br>(7.36–11.3 million) | 4.19 million<br>(3.64–5.55 million) | 90,700<br>(70,100–131,000) | 5,550<br>(3,540–10,500) |
| All influenza | 20.9 million<br>(18.9–27.0 million) | 10.6 million<br>(9.54–13.6 million) | 289,000<br>(233,000–439,000) | 19,400<br>(13,100–39,000) |

## Discussion

In this paper, we have demonstrated an extension to current CDC methods for estimating disease burden and burden prevented by vaccination for influenza that partitions burden among different influenza virus types and subtypes. During the 2016/17 to 2019/20 seasons, we estimated substantial influenza-associated disease burden and burden prevented by vaccination. In general, influenza A viruses were the greatest contributor to disease burden, particularly A(H3N2), and children <18 years were more impacted by influenza B viruses than other age groups. The greatest percentage of hospitalizations were prevented by vaccination against influenza A(H1N1) and B viruses. Overall, our framework can improve understanding of the impact of influenza each season and enhance communications of the benefits of influenza vaccination.

There are many factors that can affect disease burden and vaccine-prevented burden each season, including virus evolution and strain circulation dynamics, VE and vaccine coverage, population immunity from prior seasons, prevalence of underlying health conditions, and other factors influencing transmission dynamics (for example, individual behavior). Lower VE for A(H3N2) viruses compared to A(H1N1) and B viruses may explain why we estimated the greatest disease burden and lowest percent of burden prevented by vaccination for A(H3N2). Similarly, a combination of relatively high vaccine coverage and VE against A(H1N1) among older age groups in 2019/20 may partly explain the high levels of vaccine-prevented burden estimated in that season. More generally, our framework provides a path to explore how these interacting factors combine to impact the burden of influenza and impact of vaccination at the population-level.

Limitations to the CDC method for estimating burden and vaccine-prevented burden for all influenza viruses have been discussed previously^3,4^. These include caveats that the framework assumes protection from vaccination is ‘all-or-nothing’, uses one value for VE against all outcomes, and does not account for indirect protection through herd immunity or waning immunity. With respect to data inputs, the vaccination coverage information was based on self-(or parent-) assessment and may be subject to recall bias, although previous work has found good agreement between self-reported vaccination status among adults and vaccination status ascertained from medical records^24^. In addition, the FluSurv-NET surveillance system covers approximately 9% of the U.S. population and may not be representative of the population as a whole. Influenza A subtype was also imputed for approximately 55-60% of influenza A hospitalizations each season and could be subject to misclassification, although the rates of this are expected to be low^6,8^. Finally, the case-hospitalization ratio and fraction of cases that seek medical care were estimated during the 2009 H1N1 pandemic and will not reflect any changes to care-seeking and hospital admission rates that have occurred since then.

When extending the method to account for virus type and subtype there are several further limitations to consider. First, we assumed the case-hospitalization ratios and care-seeking fractions were equal for influenza A(H1N1), A(H3N2) and B. Further information on whether care-seeking and rates of hospital admission change by virus type and subtype would be needed to incorporate this into our extended framework. Secondly, by running independent models for each type and subtype, we implicitly assumed that an individual could be infected by multiple viruses within the same season. Although this assumption ignores any cross-immunity between A(H1N1), A(H3N2), or B viruses, we expect the individual-level risk of being infected by more than one influenza virus to be relatively low within a season and thus our results should not be substantially impacted. Finally, we did not partition estimates of disease burden or vaccine-prevented burden by influenza B lineages (B/Yamagata and B/Victoria) as the hospitalization and outpatient data did not contain this information. We have explored additional methods to partition estimates by influenza B lineage in related work^25^.

Here we have demonstrated how existing methods to estimate the disease burden and vaccine-prevented burden of influenza can be extended to estimate contributions from different virus types and subtypes. We found substantial disease burden and vaccine-prevented burden for all influenza types and subtypes across seasons and age groups, underscoring the importance of annual influenza vaccination. This is particularly relevant given recent declines in influenza vaccine coverage since the COVID-19 pandemic^12^. In addition, our results included complex variability by season, age, and vaccine coverage and effectiveness, and can be used to track changes in influenza strain dynamics in future seasons while enhancing our understanding of the burden of influenza viruses and impact of vaccination.

## Acknowledgments

The authors thank Anup Srivastav, Anurag Jain, and Mei-Chuan Hung for their contributions to obtaining the vaccination coverage estimates.

## Author Contributions

All authors attest they meet the ICMJE criteria for authorship.

## Data availability statement

All data were the result of secondary analyses and used solely as model inputs. The majority are publicly available, and the relevant links are cited in the text. A small portion are not publicly available due to privacy restrictions but are available upon reasonable request to the authors.

