## Supplementary Information for "Estimating historical disease burden and the impact of vaccination by influenza type and subtype in the United States, 2016-2020"

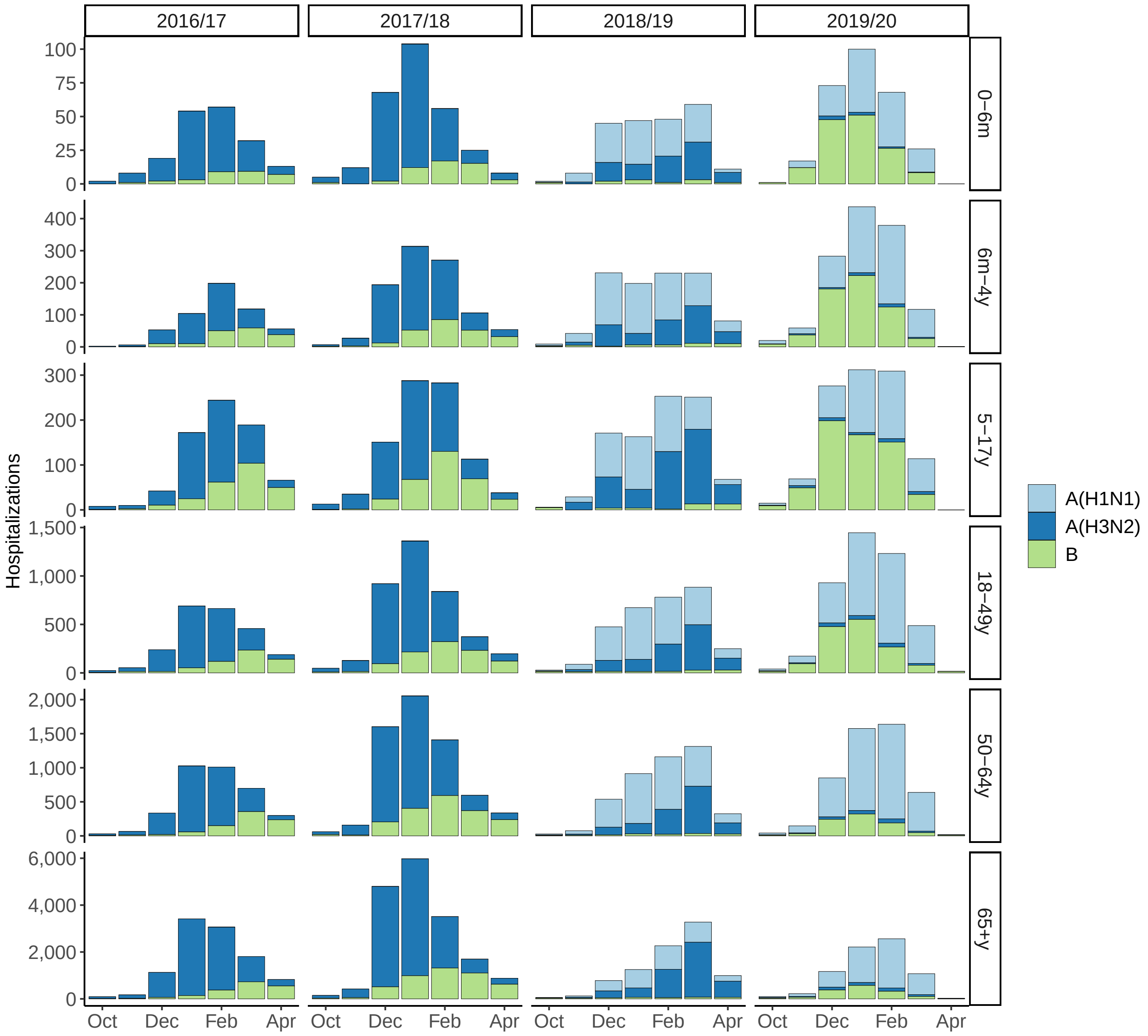


**Figure S1 – Monthly reported FluSurv-NET hospitalizations by age and virus type or subtype.**

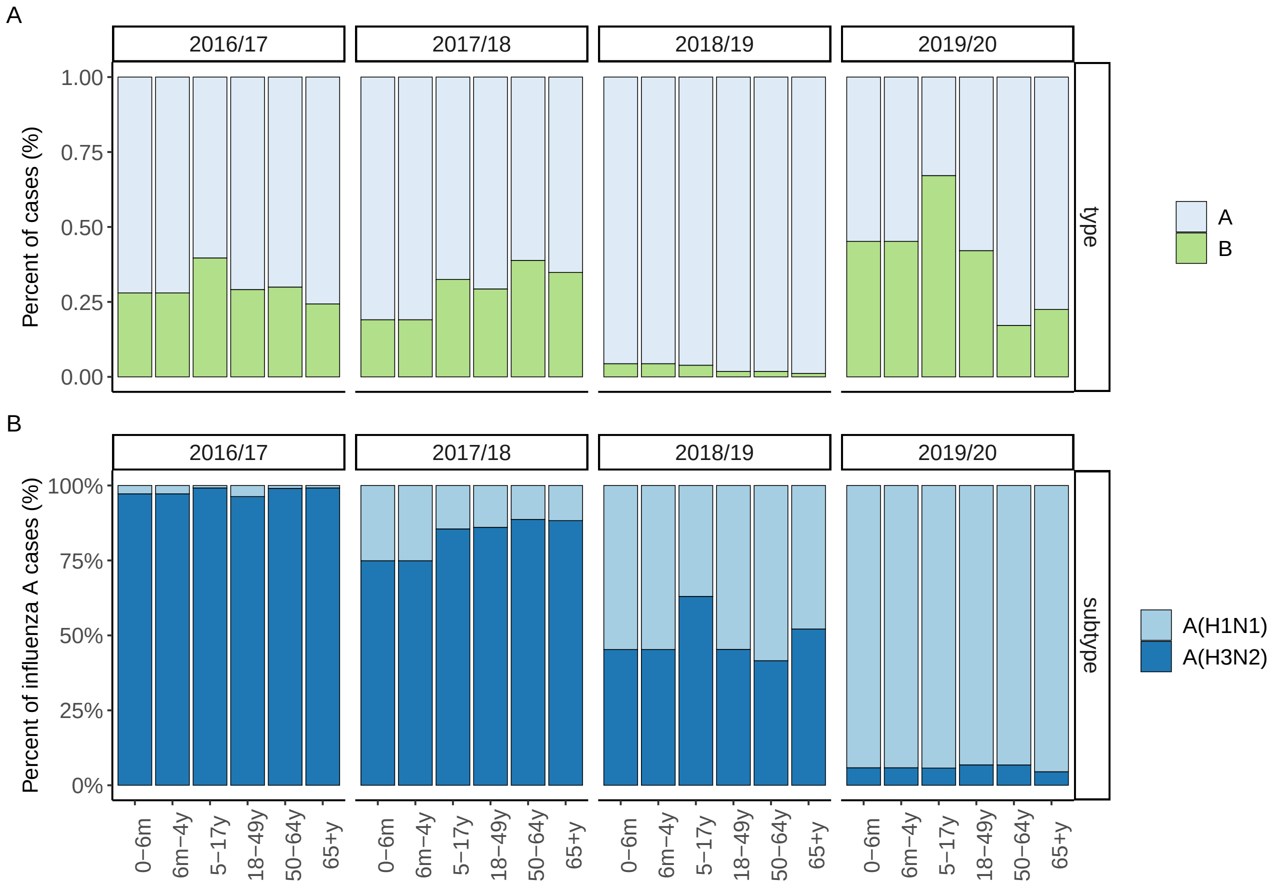


**Figure S2 – Distribution of virus type and subtypes among Flu VE Network cases.** (A) Distribution of influenza types A and B. (B) Distribution of influenza A subtypes, A(H1N1) and A(H3N2).

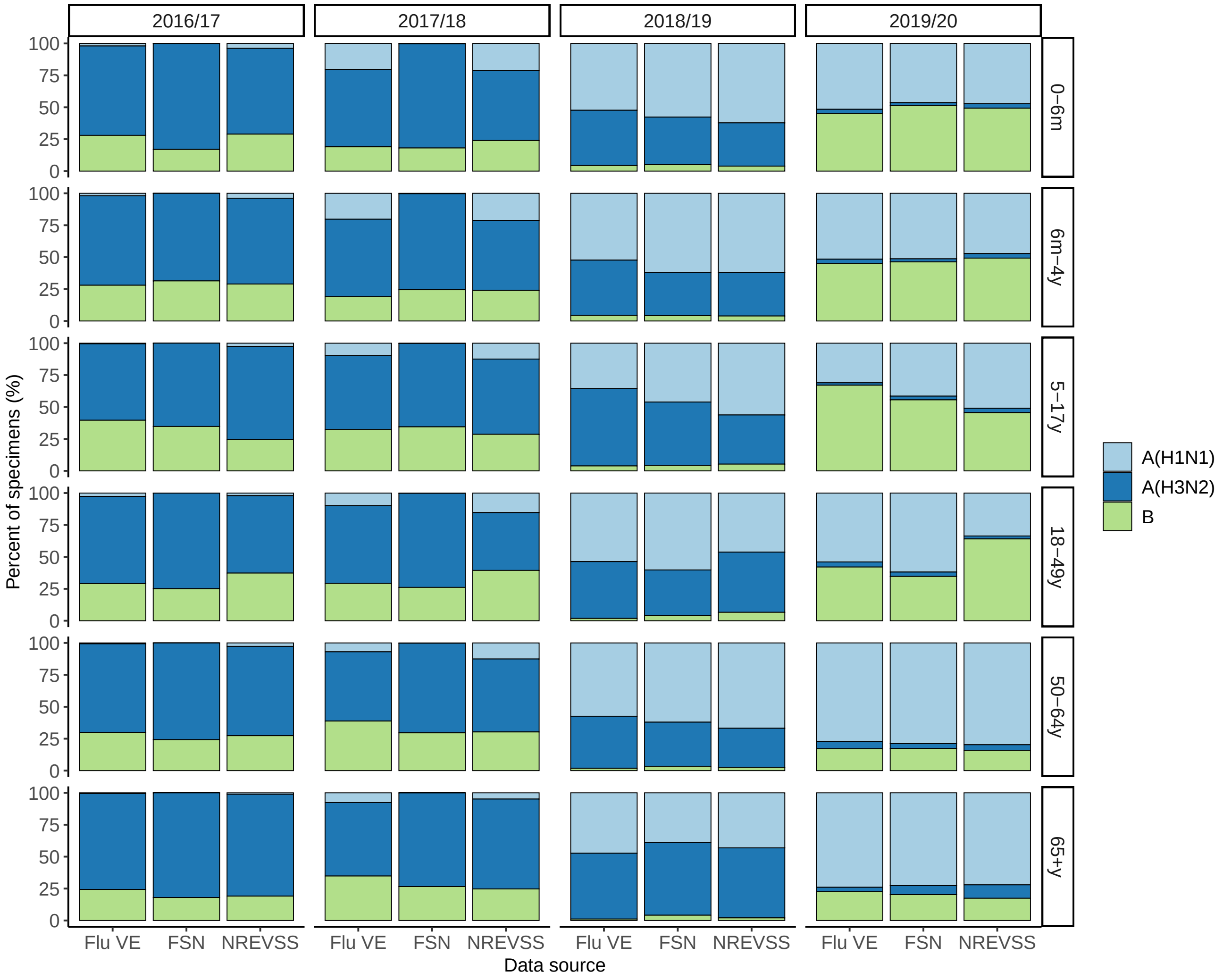


**Figure S3 – Comparison of the distribution of virus type and subtypes among independent data sources.** Flu VE refers to the Flu VE Network; FSN refers to FluSurv-NET; and NREVSS refers to the World Health Organization (WHO) and National Respiratory and Enteric Virus Surveillance System (NREVSS) Collaborating Labs. The NREVSS data were available for five age groups: 0–4 years, 5–17 years, 18–49 years, 50–64 years, and $\geq$65 years. NREVSS distributions among children 0–4 years were compared to FSN and Flu VE distributions among children 0–6 months and among 6 months–4 years.

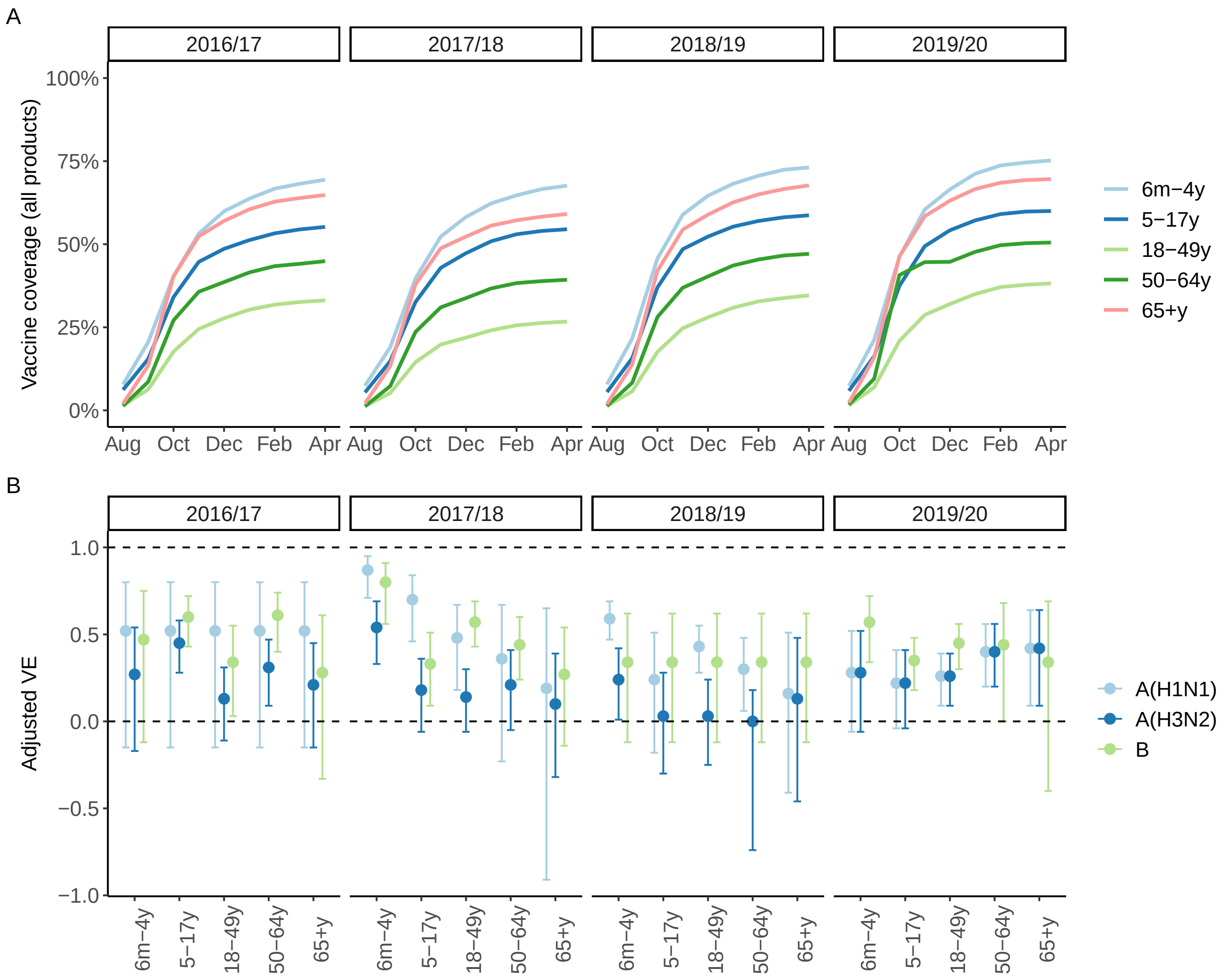


**Figure S4 – Vaccine coverage and vaccine effectiveness (VE).** (A) Monthly vaccine coverage estimates from FluVaxView. (B) VE estimates from the Flu VE Network.

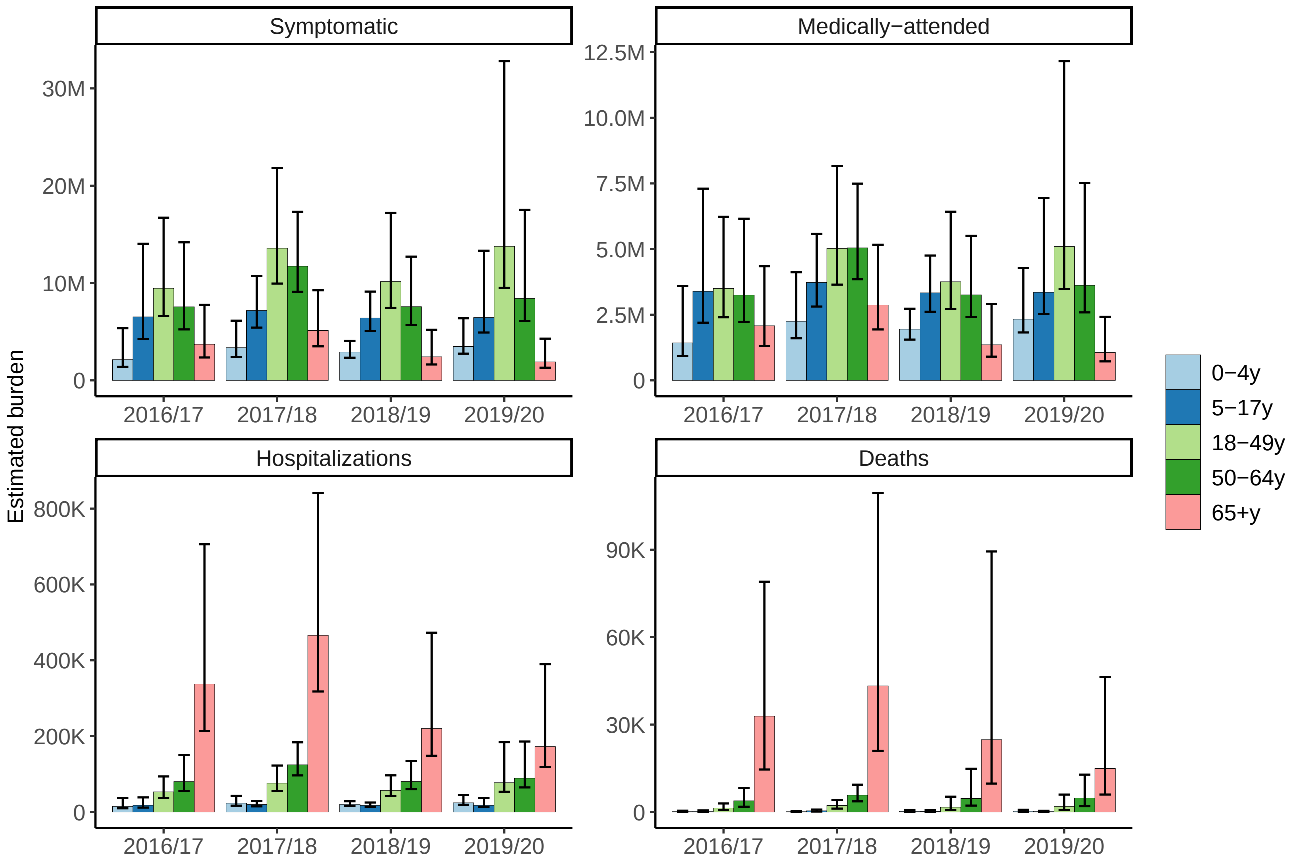


**Figure S5 – Estimated disease burden by season and age group for all influenza viruses.** Bars represent point estimates and error bars are the 95th percentile uncertainty intervals.

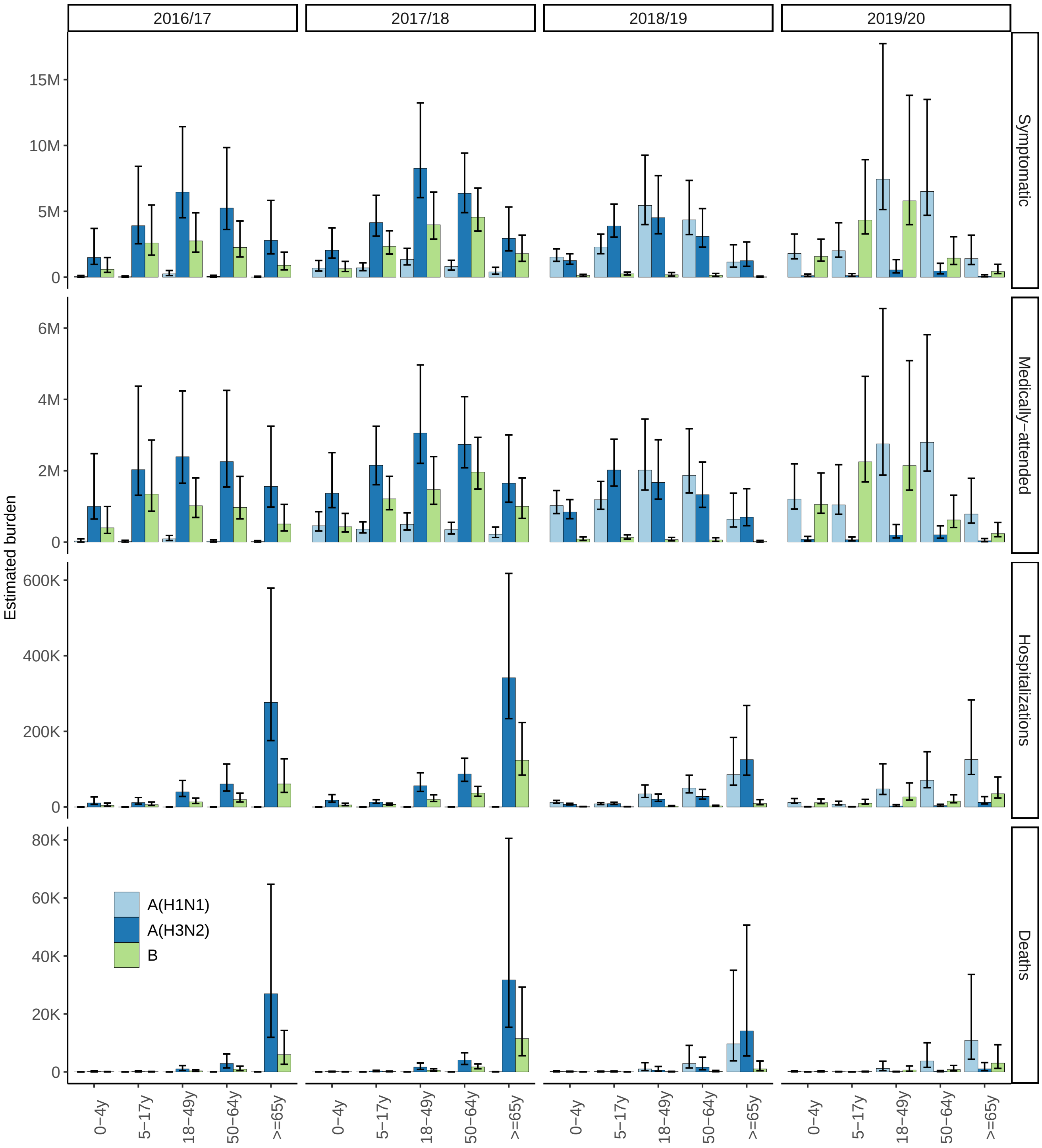


**Figure S6 – Estimated disease burden by season, age group, and virus type or subtype.** Bars represent point estimates and error bars are the 95th percentile uncertainty intervals.

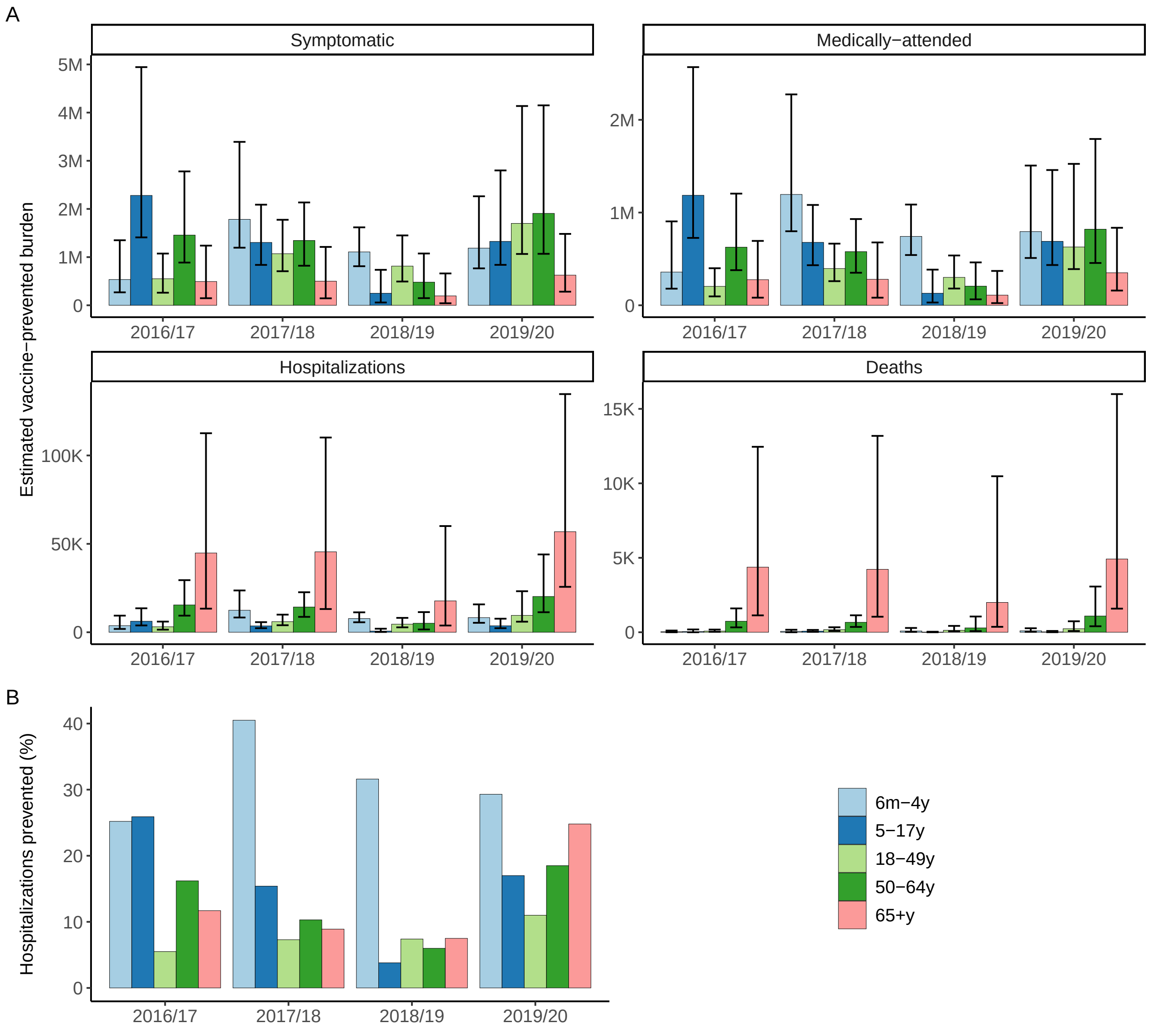


**Figure S7 – Estimated vaccine-prevented burden by season and age group for all influenza viruses.** (A) Absolute disease burden prevented by vaccination. Bars represent point estimates and error bars are the 95th percentile uncertainty intervals. (B) Hospitalizations prevented as a percentage of the total number of hospitalizations estimated to occur in the absence of vaccination.

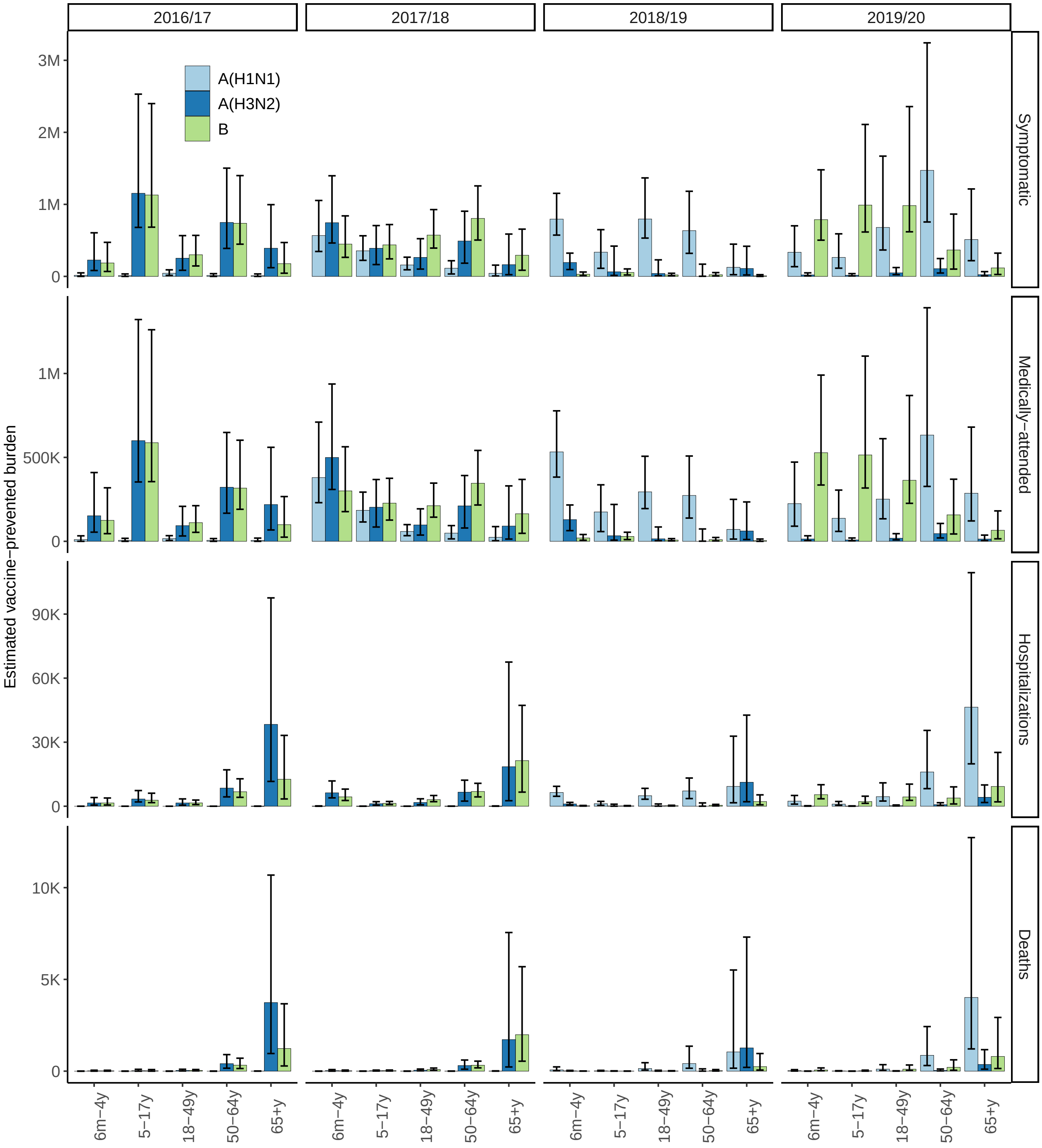


**Figure S8 – Estimated vaccine-prevented burden by season, age group, and virus type or subtype.** Bars represent point estimates and error bars are the 95th percentile uncertainty intervals.

**Table S1** – Summary values from select data sources. Values represent ranges for all influenza viruses from 2016/17–2019/20. Sources are cited in the text. Abbreviations: FSN = FluSurvNET.

| Age (years) | FSN total reported hospitalizations | Final vaccine coverage (%) | Vaccine effectiveness |
|---|---|---|---|
| 0-6m | 185 - 285 | | |
| 6m-4y | 537 - 1296 | 67.6 - 75.2 | 0.4 - 0.68 |
| 5-17y | 731 - 1095 | 54.5 - 60 | 0.07 - 0.51 |
| 18-49y | 2316 - 4326 | 26.7 - 38.2 | 0.19 - 0.34 |
| 50-64y | 3463 - 6222 | 39.3 - 50.5 | 0.14 - 0.4 |
| 65+y | 7366 - 17433 | 59.1 - 69.6 | 0.12 - 0.39 |

**Table S2** – Seasonal distribution of virus types and subtypes among all outpatient cases (from the Flu VE Network) and all hospitalizations (from FluSurv-NET).

| Season | Source | A(H1N1) (%) | A(H3N2) (%) | B (%) |
|---|---|---|---|---|
| 2016/17 | Hospitalizations | 0.01 - 0.05 | 65.23 - 82.98 | 16.97 - 34.76 |
| 2016/17 | Outpatient cases | 0.49 - 2.63 | 59.87 - 75.08 | 24.32 - 39.64 |
| 2017/18 | Hospitalizations | 0.07 - 0.3 | 65.3 - 81.6 | 18.13 - 34.5 |
| 2017/18 | Outpatient cases | 6.93 - 20.32 | 54.25 - 60.84 | 19.03 - 38.82 |
| 2018/19 | Hospitalizations | 38.98 - 62.06 | 33.9 - 56.88 | 3.42 - 5.04 |
| 2018/19 | Outpatient cases | 35.58 - 57.4 | 40.81 - 60.56 | 1.15 - 4.36 |
| 2019/20 | Hospitalizations | 41.49 - 78.84 | 2.29 - 7.04 | 17.38 - 55.62 |
| 2019/20 | Outpatient cases | 31.01 - 77.25 | 1.89 - 5.62 | 17.13 - 67.1 |

**Table S3 – Input parameters that remain constant by month and by virus type and subtype.** For parameters that vary by season, the range of inputs for 2016/17–2019/20 is shown. Sources for these values are cited in the text. Abbreviations: CHR = case-hospitalization ratio; DHR = death-hospitalization ratio; MA = medically-attended fraction.

| Age (years) | Under-detection multiplier | DHR | CHR | MA |
|---|---|---|---|---|
| 0–4 | 1.357–1.804 | 0.005–0.012 | 143.4 | 0.670 |
| 5–17 | 1.418–2.121 | 0.007–0.021 | 364.7 | 0.520 |
| 18–49 | 1.680–2.104 | 0.025–0.029 | 178.2 | 0.370 |
| 50–64 | 1.586–1.988 | 0.047–0.058 | 94.3 | 0.430 |
| ≥65 | 1.852–2.470 | 0.087–0.113 | 11.0 | 0.560 |

**Table S4 – Monte Carlo simulation settings.** Parameter notation is as follows: $\lambda$ reflects the mean of a Poisson distribution; $N$ and $p$ reflect the number of trials and probability of success of a Binomial distribution; $\mu$ and $\sigma$ reflect the mean and standard deviation of a Normal distribution; and $m$, $m_l$ and $m_u$ reflect the mode, minimum and maximum of a Beta-PERT distribution. Normal distributions were truncated at 0. Abbreviations: VE = vaccine effectiveness.

| Input | Stratifications | Distribution | Parameter(s) |
|---|---|---|---|
| Number of FluSurv-NET hospitalizations* | Season, month, age | | |
| A(H1N1) | | Poisson | $\lambda$ = observed number |
| A(H3N2) | | Poisson | $\lambda$ = observed number |
| B | | Poisson | $\lambda$ = observed number |
| Number of Flu VE Network outpatient visits* | Season, month, age | | |
| A(H1N1) | | Binomial | $N$ = observed number of influenza A visits |
| | | | $p$ = observed proportion of A visits that were A(H1N1) |
| A(H3N2) | | Binomial | $N$ = observed number of influenza A visits |
| | | | $p$ = observed proportion of A visits that were A(H3N2) |
| B | | Binomial | $N$ = observed total number of influenza visits |
| | | | $p$ = observed proportion of total visits that were B |
| Multipliers | | | |
| Under-detection of hospitalizations | Season, age | Beta-PERT | $m$ = observed value |
| | | | $m_l$ = observed lower 95th percentile |
| | | | $m_l$ = observed upper 95th percentile |
| Medically-attended fraction | Season, age | Normal | $\mu$ = observed fraction |
| | | | $\sigma$ = observed standard error |
| Death-hospitalization ratio | Season, age | See [1] | |
| Vaccination | | | |
| Coverage | Season, month, age | Normal | $\mu$ = observed fraction |
| | | | $\sigma$ = observed standard error |
| logarithm of 1 - VE | Season, month, age, type/subtype | Normal | $\mu$ = observed log(1 - VE) |
| | | | $\sigma$ = observed standard error of log(1 - VE) |

*Total hospitalizations (or outpatient visits) were calculated as the sum of simulated hospitalizations (or outpatient visits) for A(H1N1) + A(H3N2) + B.
